# A multi-centre service evaluation of the impact of the COVID-19 pandemic on presentation of newly diagnosed cancers and type 1 diabetes in children in the UK

**DOI:** 10.1101/2021.02.09.21251149

**Authors:** COVID-19 Pandemic UK-based Interest Group of Childhood Cancer and Diabetes, Gemma Williams, Ross McLean, Jo-Fen Liu, Timothy Ritzmann, Madhumita Dandapani, Dhurgshaarna Shanmugavadivel, Pooja Sachdev, Mark Brougham, Rod Mitchell, Nicholas T Conway, James Law, Alice Cunnington, Gbemi Ogunnaike, Karen Brougham, Elizabeth Bayman, David A Walker

**Author notes:** **Corresponding author**: Professor David Walker, Children’s Brain Tumour Research Centre, University of Nottingham, Nottingham NG7 2UH.

## Abstract

**Background:** The COVID-19 pandemic led to changes in patterns of presentation to Emergency Departments (ED). Child health professionals were concerned that this could contribute to the delayed diagnosis of life-threatening conditions, including childhood cancer (CC) and type 1 diabetes (T1DM). Our multicentre, UK-based service evaluation assessed diagnostic intervals and disease severity for these conditions.

**Methods:** We collected presentation route, timing and disease severity for children with newly diagnosed CC in three principal treatment centres, and T1DM in four centres between 1^st^January – 31^st^ July 2020 and the corresponding period in 2019. We assessed the impact of lockdown on total diagnostic interval (TDI), patient interval (PI), system interval (SI) and disease severity.

**Findings:** For CCs and T1DM, the route to diagnosis and severity of illness at presentation were unchanged across all time periods. Diagnostic intervals for CCs during lockdown were comparable to that in 2019 (TDI 4.6, PI 1.1 and SI 2.1 weeks), except for an increased PI in Jan-Mar 2020 (median 2.7 weeks). Diagnostic intervals for T1DM during lockdown were similar to that in 2019 (TDI 16 vs 15 and PI 14 vs 14 days), except for an increased PI in Jan-Mar 2020 (median 21 days).

**Interpretation:** There is no evidence of diagnostic delay or increased illness severity for CC or T1DM, during the first phase of the pandemic across the participating centres. This provides reassuring data for children and families with these life-changing conditions.

**Research in Context:** *Evidence before this study:* This project was initiated after the first national lockdown in March 2020 during COVID-19 pandemic in the UK. At the design stage, Medline was searched (with no language limit), using the keywords ((Cancer) OR (neoplasm) OR (Type 1 diabetes mellitus)) AND ((Covid-19) OR (SARS-CoV-2) OR (Pandemics)) AND ((Emergency department attendances) OR (diabetes ketoacidosis) OR (Delayed diagnosis) OR (interval) OR (wait)) to identify publications reporting the impact of the pandemic and public health measures on both overall and paediatric healthcare services. Significant changes in service utilisation in the UK were reported following the commencement of the first lockdown, including a 49% reduction in emergency department attendances in the week following the lockdown; and two adult studies reported that referral via the urgent two-week wait cancer referral diagnoses decreased by 84% from Mar-May and 60% in June 2020. As for Type 1 diabetes (T1DM), a 30 patient UK-study reported an increase in newly diagnosed T1DM during the first six weeks of lockdown. Increased proportions of severe diabetic ketoacidosis (DKA) at presentation were also reported in an Italian survey involving 53 paediatric diabetes centres. Through the search we identified a need for multi-centre, more thorough assessment on referral pathways, time taken from symptom onset to diagnosis, and its association with severity at presentation for children diagnosed with life-changing conditions during the national lockdown.

*Added value of this study:* Our findings suggest that the first national lockdown in the UK were not associated with delayed diagnosis of childhood cancer or type 1 diabetes at participating centres. This provides reassuring information for children and families with these life-changing conditions.

*Implication:* We believe that our study can play a key role in allaying parental and professional concern. it is important to establish whether subsequent public health measures have impacted the diagnostic interval in the context of an evolving backlog of patient referrals across the UK.

## INTRODUCTION

The UK public health response to the COVID-19 pandemic led to major changes in service utilisation of Emergency Departments (ED), with an observed 49% reduction in attendances in the week following the March 2020 announcement of lockdown. ^1^ More specifically, this reduction was also seen in children’s ED attendances in both Scotland and Italy ^2,3^. This raised concern amongst child health professionals that the pandemic could be associated with delayed presentation of children with significant illnesses ^4^ including life-changing childhood conditions such as type 1 diabetes (T1DM) and childhood cancer (CC). We were concerned there may have been an increase in delays in seeking medical advice (the patient interval – PI) and/or accessing initial investigations and interventions (the system interval – SI), leading to a prolongation of the total diagnostic interval (TDI).^5^

In the UK, 1900 children are diagnosed with cancer each year ^6,7^. The diverse and often insidious nature of presenting symptoms, combined with the low incidence and lack of awareness of childhood malignancy may contribute to a prolongation of the TDI. Recent adult studies ^8,9^ report that referrals via the urgent two-week wait pathway for suspected cancer diagnoses decreased by 84% from March-May 2020 ^9^and 60% in June 2020.^8^ One study predicted a reduction of over 10% in 10 year survival of adults with cancer^9^ whilst another predicted an excess of 1307 cancer deaths.^8^ The evidence is equivocal as to whether delays in time to diagnosis impact upon survival from CCs ^9-11^, however the impact of psychological and economic distress on families awaiting a CC diagnosis should not be. underestimated ^10^

Experience of the COVID-19 pandemic for children with T1DM has been inconsistent. A survey of 53 Italian paediatric diabetes centres found that the number of children with a new diagnosis of diabetes from February-April 2020 was 23% lower than the corresponding period in 2019. The survey also found the proportion of patients presenting with severe diabetic ketoacidosis (DKA) was increased ^12^. A 30 patient UK-study reported an apparent increase in new-onset T1DM during the first six weeks of lockdown ^13^.

The aim of the project was to assess any impact of the COVID-19 pandemic on new diagnoses of CC and T1DM at participating UK centres.

## METHODS

### Study Design

We undertook a multicentre service evaluation using existing clinical case data assessing the route of diagnosis, diagnostic interval and severity of presentation.

#### Eligibility criteria were

1. All children who attended three centres (the Royal Hospital for Sick Children Edinburgh, Leeds Teaching Hospitals NHS Trust and Nottingham University Hospitals NHS Trust) and were diagnosed with cancer between 1^st^ January and 30^th^ June 2019 and the corresponding period in 2020;
2. All children who attended four centres (the Royal Hospital for Sick Children, Edinburgh (RHSC), Nottingham University Hospital NHS Trust (NUH), University Hospital Wishaw (UHW) and Ninewells Hospital, Dundee and were diagnosed with T1DM between 1^st^ January-31^st^ July 2019 and the corresponding period in 2020.

Whilst analyses were prospectively designed, a standard proforma (online supplementary material) was used to retrospectively collect information on demographics, diagnosis, referral pathway and clinical presentation. We also collected information on whether patients were shielding at presentation (following specific government guidelines to minimise risk of SARS-CoV-2 exposure for those considered clinically extremely vulnerable). Dates of symptom onset, first presentation to healthcare and final diagnosis were used to calculate total diagnostic interval (TDI, time between symptom onset to diagnosis), patient interval (PI, time between symptom onset to first presentation) and system interval (SI, time between first presentation to diagnosis)^5^

The protocol was approved by Caldicott Guardians of all centres involved.

### Statistical analysis

Descriptive analyses, *χ*^2^ test, Mann-Whitney U or Kruskal-Wallis tests were used to describe patterns of referral and illness, comparing the differences of key measures among different time periods (1^st^ January −31^st^ March 2020, and 1^st^ April-31^st^ July 2020, and the corresponding period in 2019). Pair-wise comparisons of proportions were carried out using the Z test with Bonferroni corrections. All analyses were performed with IBM SPSS 26.0 for Windows (IBM Corp. Armonk, NY, USA) and p<0.05 was considered statistically significant.

## RESULTS

### Study population

#### Childhood Cancer

There were 253 new diagnoses of childhood cancer during the study period (Table 1). Of these, 164 (64%) were male and 55 (22%) were from a BAME background. Patients were diagnosed at one of three principal treatment centres (Edinburgh=64, Leeds=100, Nottingham=89). There were no differences in gender, ethnic background or age at diagnosis between study periods (Table 1). The proportion of tumour type in each evaluation period did not change. 95% (53/56) of those who presented during the lockdown period were not shielding.

**Table 1.** Summary of incident childhood cancer patient characteristics (n=253)

|  | **Total**<br>**(n=253)** |  | **Jan -Jun**<br>**2019**<br>**(n=138)** |  | **Jan-Mar**<br>**2020**<br>**(n=59)** |  | **Apr-Jun**<br>**2020**<br>**(n=56)** |  | **p-value** |
| --- | --- | --- | --- | --- | --- | --- | --- | --- | --- |
|  | n | CoI% | n | CoI% | n | CoI% | n | CoI% |  |
| **Gender** |  |  |  |  |  |  |  |  | 0.150 |
| Male | 162 | 64% | 81 | 59% | 41 | 69% | 40 | 71% |  |
| Female | 91 | 36% | 57 | 41% | 18 | 31% | 16 | 29% |  |
| **Age** |  |  |  |  |  |  |  |  | 0.963 |
| Under 5 | 99 | 39% | 53 | 38% | 25 | 43% | 21 | 38% |  |
| 5-11 | 92 | 37% | 52 | 38% | 19 | 33% | 21 | 38% |  |
| 12+ | 61 | 24% | 33 | 24% | 14 | 24% | 14 | 25% |  |
| **BAME background** |  |  |  |  |  |  |  |  | 0.945 |
| No | 198 | 78% | 107 | 78% | 47 | 80% | 44 | 79% |  |
| Yes | 55 | 22% | 31 | 22% | 12 | 20% | 12 | 21% |  |
| **COVID isolation/shielding** |  |  |  |  |  |  |  |  |  |
| No | 250 | 99% | 138 | 100% | 59 | 100% | 53 | 95% |  |
| Yes | 3 | 1% | 0 | 0% | 0 | 0% | 3 | 5% |  |
| **Tumour type** |  |  |  |  |  |  |  |  | 0.273 |
| Leukaemia | 69 | 27% | 33 | 24% | 17 | 29% | 19 | 34% |  |
| CNS tumour | 74 | 29% | 48 | 35% | 14 | 24% | 12 | 21% |  |
| All other tumour types | 110 | 43% | 57 | 41% | 28 | 47% | 25 | 45% |  |
| **ICU stay** |  |  |  |  |  |  |  |  | 0.275 |
| No | 223 | 88% | 120 | 87% | 55 | 93% | 48 | 86% |  |
| Yes | 26 | 10% | 17 | 12% | 3 | 5% | 6 | 11% |  |
| Not known | 4 | 2% | 1 | 1% | 1 | 2% | 2 | 4% |  |
| **HCP visits before diagnosis** |  |  |  |  |  |  |  |  | 0.569 |
| 3 or less | 152 | 60% | 82 | 59% | 40 | 68% | 30 | 54% |  |
| 4-6 | 64 | 25% | 38 | 28% | 12 | 20% | 14 | 25% |  |
| 7-9 | 21 | 8% | 11 | 8% | 3 | 5% | 7 | 13% |  |
| 10 or more | 16 | 6% | 7 | 5% | 4 | 7% | 5 | 9% |  |

Overall there was a 17% reduction in number of incident childhood cancer cases between 2019 (n=138) and 2020 (n=115). This change varied between centres (4% increase to 40% reduction) (Figure 1).

**Figure 1.**
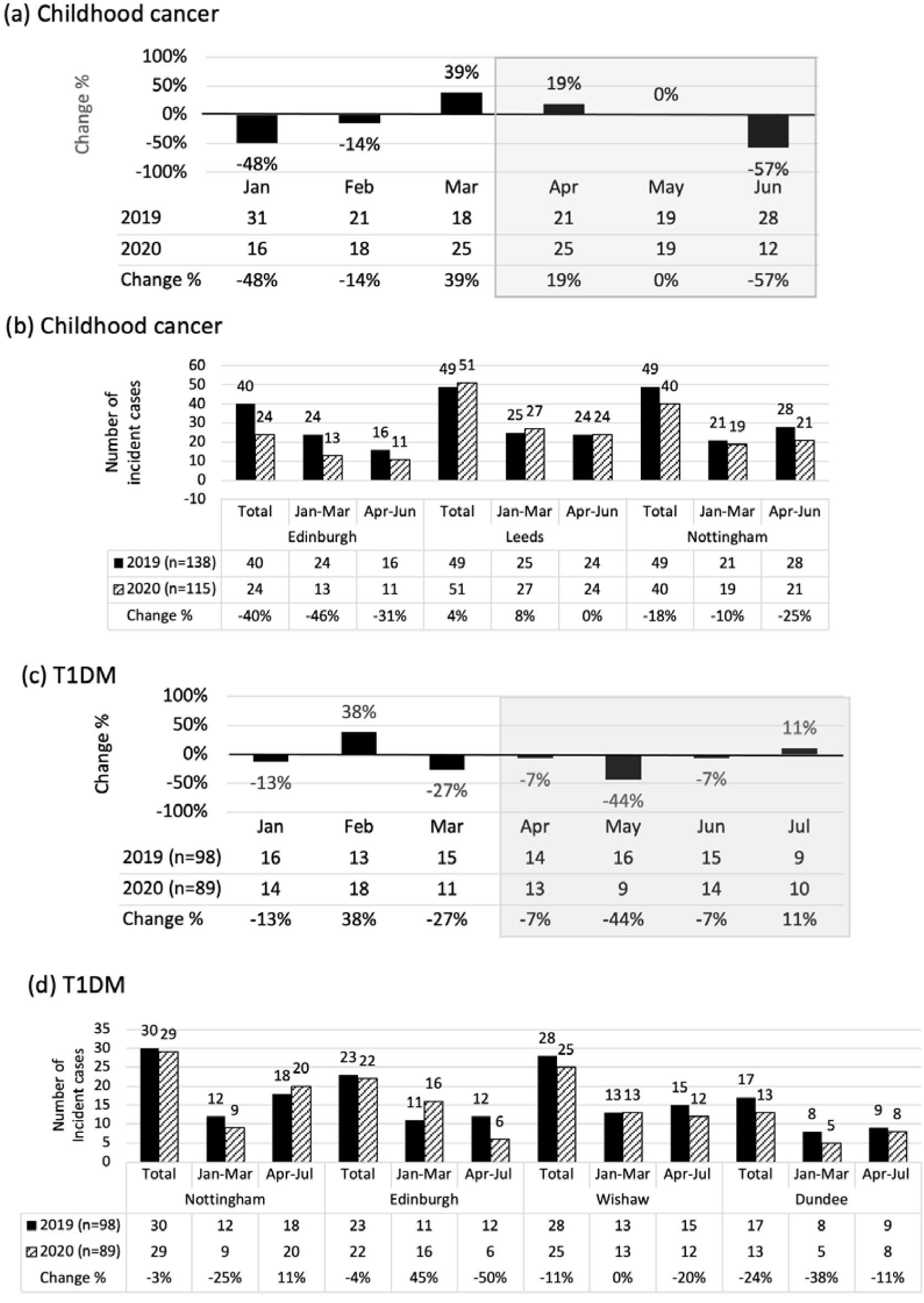
Number of newly diagnosed of childhood cancer and T1DM cases between Jan-Jul 2020 compared to the corresponding period in 2019. **(a) (c)** data covers all participating centres contributing data to the project by month. Shaded areas indicate national lockdown months **(b) (d)** data by individual centre over the periods January to June/July in 2019 and 2020.

#### Type 1 Diabetes

There were 187 new diagnoses of T1DM during the study period (Table 2). Of these, 90 (48%) were male and 18 (10%) were from a BAME background. Patients were diagnosed at one of four participating centres (Edinburgh=45, Dundee=30, Wishaw=53, Nottingham=59). There were no differences in gender, ethnic background or age at diagnosis between the study periods (Table 2). 91% (42/46) of those who presented during the lockdown period were not shielding.

**Table 2.** Summary of incident TIDM patient characteristics (n=187)

|  | Total<br>(n=187) |  | Jan-Jul<br>2019<br>(n=98) |  | Jan-Mar<br>2020<br>(n=43) |  | Apr-Jul<br>2020<br>(n=46) |  | p-value |
| --- | --- | --- | --- | --- | --- | --- | --- | --- | --- |
|  | n | Col% | n | Col% | n | Col% | n | Col% |  |
| **Gender** |  |  |  |  |  |  |  |  | 0.718 |
| Male | 90 | 48% | 46 | 47% | 23 | 53% | 21 | 46% |  |
| Female | 97 | 52% | 52 | 53% | 20 | 47% | 25 | 54% |  |
| **Age** |  |  |  |  |  |  |  |  | 0.174 |
| Under 5 | 32 | 17% | 22 | 22% | 5 | 12% | 5 | 11% |  |
| 5-11 | 87 | 47% | 43 | 44% | 18 | 42% | 26 | 57% |  |
| 12+ | 68 | 36% | 33 | 34% | 20 | 47% | 15 | 33% |  |
| **BAME background** |  |  |  |  |  |  |  |  | 0.484 |
| No | 168 | 90% | 87 | 89% | 41 | 95% | 40 | 89% |  |
| Yes | 18 | 10% | 11 | 11% | 2 | 5% | 5 | 11% |  |
| **COVID isolation/shielding** |  |  |  |  |  |  |  |  | 0.012 |
| No | 183 | 98% | 98 | 100% | 43 | 100% | 42 | 91% |  |
| Yes - self-isolation | 3 | 2% | 0 | 0% | 0 | 0% | 3 | 7% |  |
| Yes - shielding | 1 | 1% | 0 | 0% | 0 | 0% | 1 | 2% |  |
| **DKA** |  |  |  |  |  |  |  |  | 0.624 |
| No | 112 | 60% | 59 | 61% | 26 | 60% | 27 | 59% |  |
| Mild/Moderate | 46 | 25% | 25 | 26% | 12 | 28% | 9 | 20% |  |
| Severe | 28 | 15% | 13 | 13% | 5 | 12% | 10 | 22% |  |
| **Ventilation** |  |  |  |  |  |  |  |  | 0.607 |
| No | 182 | 97% | 94 | 96% | 43 | 100% | 45 | 98% |  |
| Yes | 5 | 3% | 4 | 4% | 0 | 0% | 1 | 2% |  |
| **ITU stay** |  |  |  |  |  |  |  |  | 0.625 |
| No | 169 | 91% | 90 | 92% | 39 | 93% | 40 | 87% |  |
| Yes | 17 | 9% | 8 | 8% | 3 | 7% | 6 | 13% |  |
| **HCP visits before diagnosis** |  |  |  |  |  |  |  |  | 0.673 |
| 1 | 164 | 92% | 87 | 92% | 37 | 95% | 40 | 89% |  |
| >1 | 15 | 8% | 8 | 8% | 2 | 5% | 5 | 11% |  |

A reduction in the numbers of new cases of T1DM between the months of Apr-Jul 2020 and the identical period in 2019 occurred in three of the four units. Overall, there was a 3-24% reduction in new diagnoses of T1DM between the months of Jan-Jul 2020 compared with the corresponding period in 2019 in all centres (Figure 1).

### Route to diagnosis

#### Childhood Cancer

Across all time points, 60% of children (152/253) were diagnosed within three or fewer healthcare contacts, 25% within four to six contacts, 8% within seven to nine contacts and 6% required more than 10 contacts prior to diagnosis. There was no difference in distribution across three time periods (p=0.569) or Jan-Mar 2020 and Apr-Jun 2020 (p=0.359). (Figure S1a). General practice was the first point of healthcare contact in the majority of patients across all time periods (54% 2019, 49% Jan-Mar 2020, 48% Apr-Jun 2020) (Figure S1b). 63% of patients presented to hospital as an emergency presentation either from Primary Care or to the ED. 36% of children had their diagnostic investigation requested as an inpatient. These proportions were consistent across all three time periods (p=0.405) (Figure S1c).

#### Type 1 Diabetes

The source of referral leading to diagnosis was the ED in 30% (14/46) of cases between Apr-Jul 2020. This compared with 12% (5/43) of cases diagnosed between Jan-Mar 2020 (p=0.091), and 19% (19/98) of cases diagnosed over the period of Jan-Jul 2019 (p=0.426). 63% of patients had been referred by their GP between Apr-Jul 2020, compared with 81% of patients in the preceding three months, and 72% of patients in 2019 (Figure S2).

### Time to diagnosis

#### Childhood Cancer

Across all three centres, there was no difference in TDI between 2019 and Jan-Mar 2020 or Apr-Jun 2020 (p=0.351) (Figure 2, Figure S3a). There was a significant increase in PI between 2019 and Jan-Mar 2020 (median 0.9 vs 2.7 weeks, p=0.005), but during Apr-Jun 2020 there was no difference compared with 2019 (median 1.1 vs 0.9 weeks, p=0.383) (Figure 2). SI was stable across all time points (Figure 2). There was no difference in TDI across all time periods for leukaemias, CNS tumours and solid tumours in subgroup analyses (Figure 3). There was no difference in PI or SI for individual principal treatment centres or tumour types (Figures 3 and Figure S3b-c).

**Figure 2.**
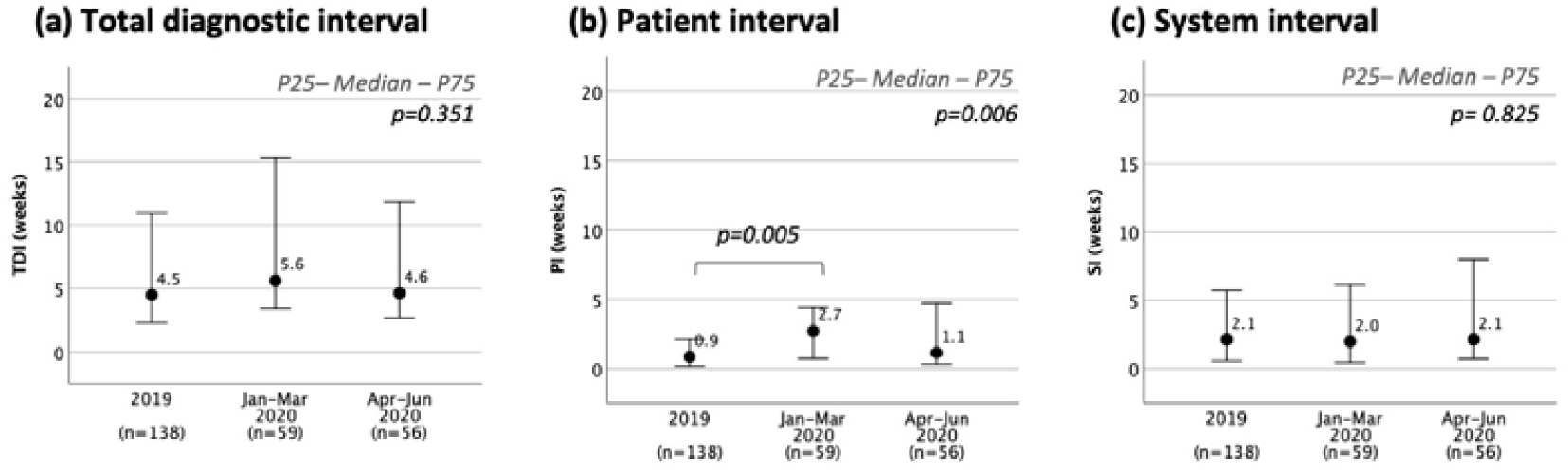
Time to diagnosis for paediatric oncology. **(a)** Total diagnostic interval (TDI): interval between first symptom onset to diagnosis. **(b)** Patient interval (PI): time from initial symptom onset to first presentation to healthcare. **(c)** System interval (SI): time between first presentation to healthcare to diagnosis

**Figure 3.**
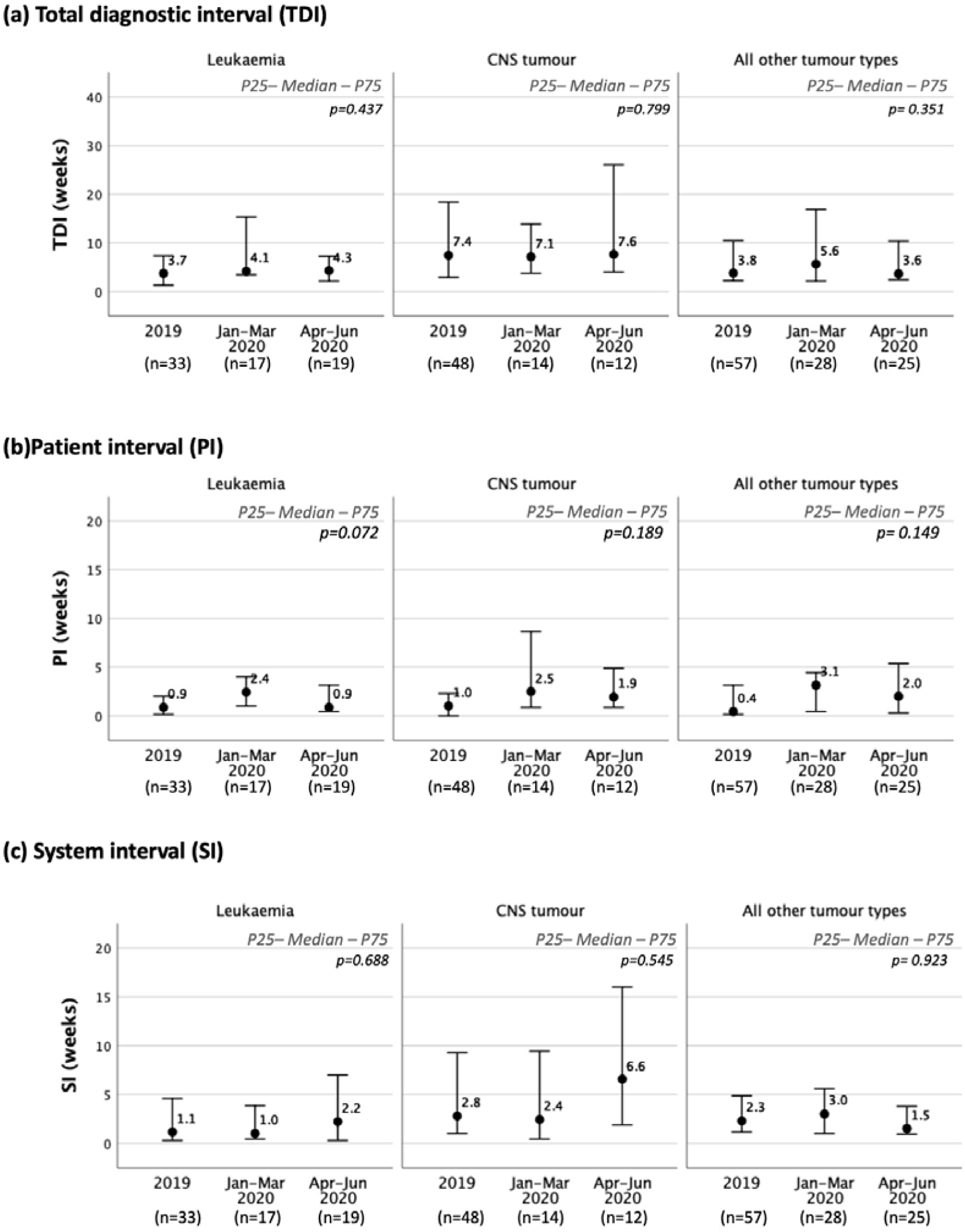
Time to diagnosis for childhood cancer for leukaemia, CNS tumour and all other tumour types combined. **(a)** Total diagnostic interval (TDI): interval between first symptom onset to diagnosis. **(b)** Patient interval (PI): time from initial symptom onset to first presentation to healthcare. **(c)** System interval (SI): time between first presentation to healthcare to diagnosis

#### Type 1 Diabetes

Median TDI for incident T1DM cases was 16, 21 and 15 days for 2019, Jan-Mar 2020 and Apr-Jul 2020 respectively. There was no difference across the three time preriods (p=0.119). A similar pattern was observed in patient interval. The comparison across all time periods was close to significance (p=0.054), sub-analyses showed that PI was longer during Jan-Mar 2020 compared to Apr-Jul 2020 (median 21 vs 14 days, p=0.025) and 2019 (median 14 days, p=0.036) (Figure 4 and Figure S4).

**Figure 4.**
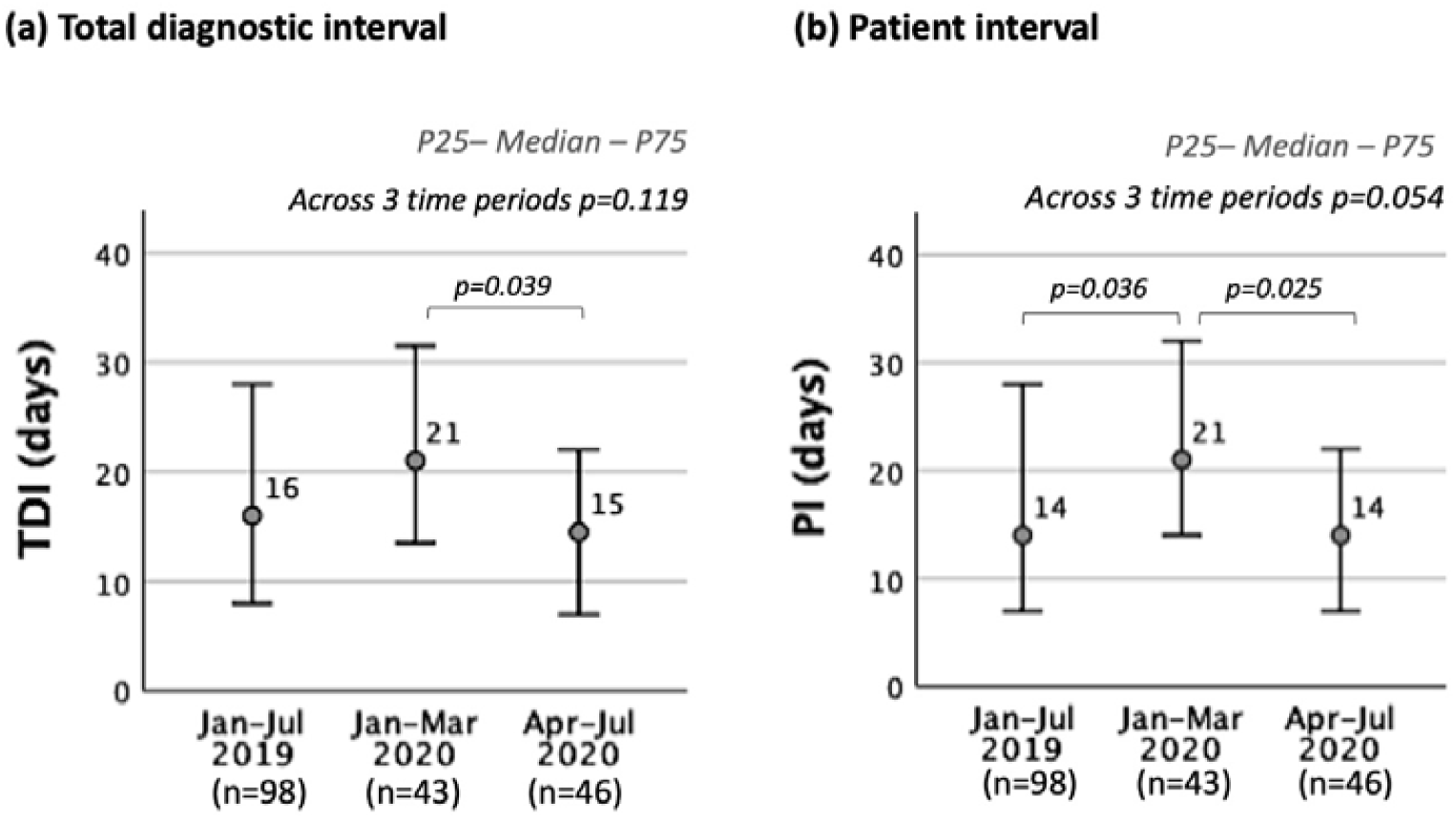
Time to diagnosis of incident T1DM cases between January and July 2020 and corresponding period in 2019. **(a)** Total diagnostic interval (TDI): interval between first symptom onset to diagnosis. **(b)** Patient interval (PI): time from initial symptom onset to first presentation to healthcare.

### Severity of Presentation

#### Childhood Cancer

Within seven days of diagnosis, 10% (26/253) of patients with childhood cancer required admission to paediatric intensive care. This was stable across all time periods (p=0.275) (Table 1).

#### Type 1 Diabetes

The proportion of patients presenting in DKA was 41% (19/46) in the period Apr-Jul 2020, 40% (17/43) for the time period Jan-Mar 2020, and 39% (38/98) for the time between Jan-Jul 2019. There was no difference in the proportion of patients in DKA across all time periods. The proportion of children presenting with severe DKA (pH<7.1, serum bicarbonate <5mmol/L) ^14^ had non-significantly increased between lockdown (22%, 10/46) compared with 12% (5/43) Jan-Mar 2020 and 13% (13/98) Jan-Jul 2019) in the periods prior to lockdown. There was no significant difference in the rate of intensive care admission or requirement for ventilatory support (Table 2).

## DISCUSSION

We have demonstrated that the first phase of the COVID-19 pandemic was not associated with adverse impact on route of presentation, TDI or disease severity at presentation for children with a new diagnosis of CC or T1DM at the study centres. Given the reported reduction in paediatric ED attendance,^15,16^ it had been predicted that both the PI and TDI would be prolonged, and clinical presentations more severe. This prediction was not supported by our study.

A snapshot survey was commissioned in April 2020 by the ChildCancerSmart Team in conjunction with the Childhood Cancer and Leukaemia Group (CCLG) to obtain the absolute numbers of new diagnoses of childhood cancer at each principal treatment centre in the UK (online supplementary material). This survey revealed 27% fewer new cases in April 2020 compared with April 2019, raising concerns about the potential for diagnostic delay. Similar anxieties had been expressed related to delayed presentation of patients with T1DM. ^17^ One survey of diabetes units in the UK reported that 20% of children and young people diagnosed with T1DM between 1 March and 30 June 2020 had had a delayed presentation. Reasons for this included fear of contracting SARS-CoV-2 as well as limited access to GP services.^13^

We demonstrated higher PI during Jan-Mar 2020 for patients with CC and T1DM. This may reflect a period of uncertainty for patients and health care systems in preparation for the first phase of the pandemic. The subsequent reduction in PI during and after lockdown suggests no significant delay in caregivers seeking medical attention for children, which may reflect contemporary public health messaging.

A study of the North-West London Paediatric Diabetes Network between 23 March and 4 June 2020 reported an apparent increase in cases of new-onset T1DM in two of five units^13^. 21/30 children with a new diagnosis presented with DKA, 52% of which were severe. These are generally considered to represent high rates both of DKA and severe DKA in newly diagnosed patients. However, the number of children involved in this study was small, no comparisons were possible with other time periods and we were unable to replicate the findings.

In a large survey of 53 Italian paediatric diabetes centres, the number of newly diagnosed children with T1DM and DKA were similar to 2019 ^12^. However the proportion of all T1DM patients who developed severe DKA was significantly greater in 2020 (44.3% v 36.1%, p=0.03)^12^. Despite not being significant, the pattern of our results are similar to the Italian experience ^12^ which demonstrated a 9% reduction in new diagnoses of T1DM when comparing Jan-Jul 2020 to 2019. Our data showed a 7-44% reduction in three of the four months following lockdown in new cases of T1DM, with a 27% decrease during March 2020 when national lockdown was announced. The incidence of DKA at presentation was stable between the measured time periods, however the incidence of severe DKA was slightly worse following lockdown (22% (Apr-Jul 2020), v 12% (Jan-Mar 2020) v 13% (Jan-Jul 2019)).

The increased incidence of severe DKA amongst patients diagnosed post-lockdown, whilst not significant, is a potential concern. This finding is consistent with the Italian data and with other units in the UK ^12,17^. It initially appears to be counter-intuitive when one takes into account the reduction in the TDI and PI post-lockdown. However, it is well recognised that some children with T1DM can present acutely with rapid onset of ketoacidosis. The increased rate of severe DKA at presentation serves to emphasise the importance of the ongoing provision of public health campaigns to raise awareness of the symptoms of T1DM amongst parents/caregivers^18^.

Overall, TDI for CCs was stable between the three time periods. Consistent with previously published evidence^11^, the TDI for leukaemia was shorter than for CNS tumours, suggesting that the COVID pandemic has had little impact on this pattern.

A large cross-sectional survey of the impact of the pandemic on CC care from the Paediatric Oncology East and Mediterranean Group, reported that some centres noticed that all newly diagnosed patients experienced delays in diagnosis.^19^ This was thought to be due to: 1) patients refusing to present for essential visits for fear of contracting COVID-19; 2) hospital staff being relocated to other areas; and 3) governmental decisions impacting upon the availability of public transport and freedom of travel. Although, in the UK during the first lockdown the availability of public transport was decreased and in some tertiary oncology centres paediatric hospital staff were redeployed, our data does not support a similar situation in the participating centres.

10% of new CC diagnoses in this study required intensive care within seven days of admission; the majority of these had CNS tumours, most likely due to the requirement for early neurosurgery. From this we infer that patients diagnosed during the pandemic were not more unwell than if they had been diagnosed earlier. We recognise that intensive care admission resulting from treatment can occur in the early stages post-diagnosis. ^20^ Consequently, using intensive care admission may overestimate initial disease severity. Reassuringly, a CCLG study reported that children with cancer and SARS-CoV-2 infection do not appear at increased risk of severe infection compared to the general paediatric population ^21^.

Whilst there was variability between units, there was evidence of a shift towards first presentations to ED from primary care, for CC and T1DM patients. Whilst we did not identify statistically significant changes, we recommend that each treatment centre evaluate any change in how their services have been accessed during the pandemic, to assist in future service planning.

There are a number of limitations to this study. Only four centres in the UK were involved, therefore the study lacked the ability to detect national variations in patterns of presentation. Whilst the analysis was prospectively designed, the data collection was retrospective. We cannot exclude the possibility of incomplete areas of data collection given that some of the children in the service evaluation will have been treated in more than one centre. We believe that this effect is both random and minimal across the centres. Given the resources available for this service evaluation we elected to collect comparison data for one year prior to the pandemic (2019). However, we recognise that fluctuation occurs and a longer period of pre-pandemic data collection would have provided greater insight into this variation. It was reassuring however that the incident cases of CC across the UK as a whole remained stable from 2013-17 ^22^.

In view of the fluid situation of the pandemic, data collection was completed in July 2020 as we believed that timely presentation could inform local practice. We will continue data collection to account for the diagnostic lag for specific diseases including brain tumours. Data collection at a more comprehensive national level would also provide greater clarity on diagnostic intervals. Furthermore, it is important to establish whether subsequent public health measures have impacted the TDI in the context of an evolving backlog of patient referrals across the UK.

## Conclusions

This project was born out of a desire to understand the impact that COVID-19 was having on life-changing childhood diagnoses. Our findings suggest that public health measures, imposed to control the spread of the pandemic during the first lockdown in the UK, were not associated with delayed diagnosis of CC or T1DM at participating centres. This is good news in the context of a pandemic that has been harmful to children’s health and wellbeing in many other ways. We believe that our study can play a key role in allaying parental and professional concern.

## Supporting information

Online supplementary material - CC data collection form

Online supplementary material - T1DM data collection form

Online supplementary material - CC cases

## Data Availability

Contact corresponding author regarding availability of data

## Acknowledgement

The authors would like to thank Dr Bob Phillips for his valuable input into the Caldicott approval application process and project set up at Leeds Teaching Hospitals NHS Trust.

