## Supplementary figures and images for "A multi-centre service evaluation of the impact of the COVID-19 pandemic on presentation of newly diagnosed cancers and type 1 diabetes in children in the UK"

### Figure S1

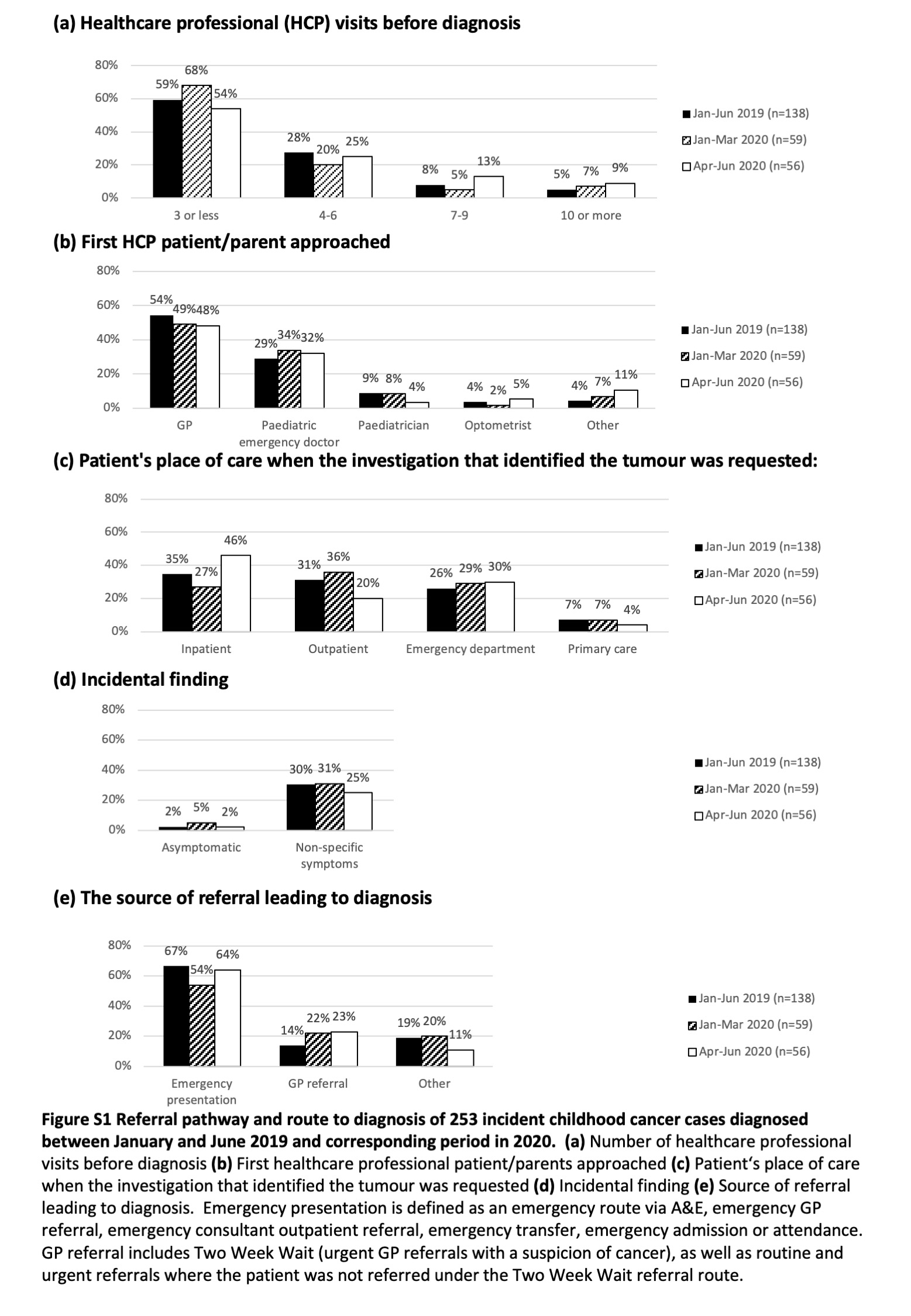

### Figure S2

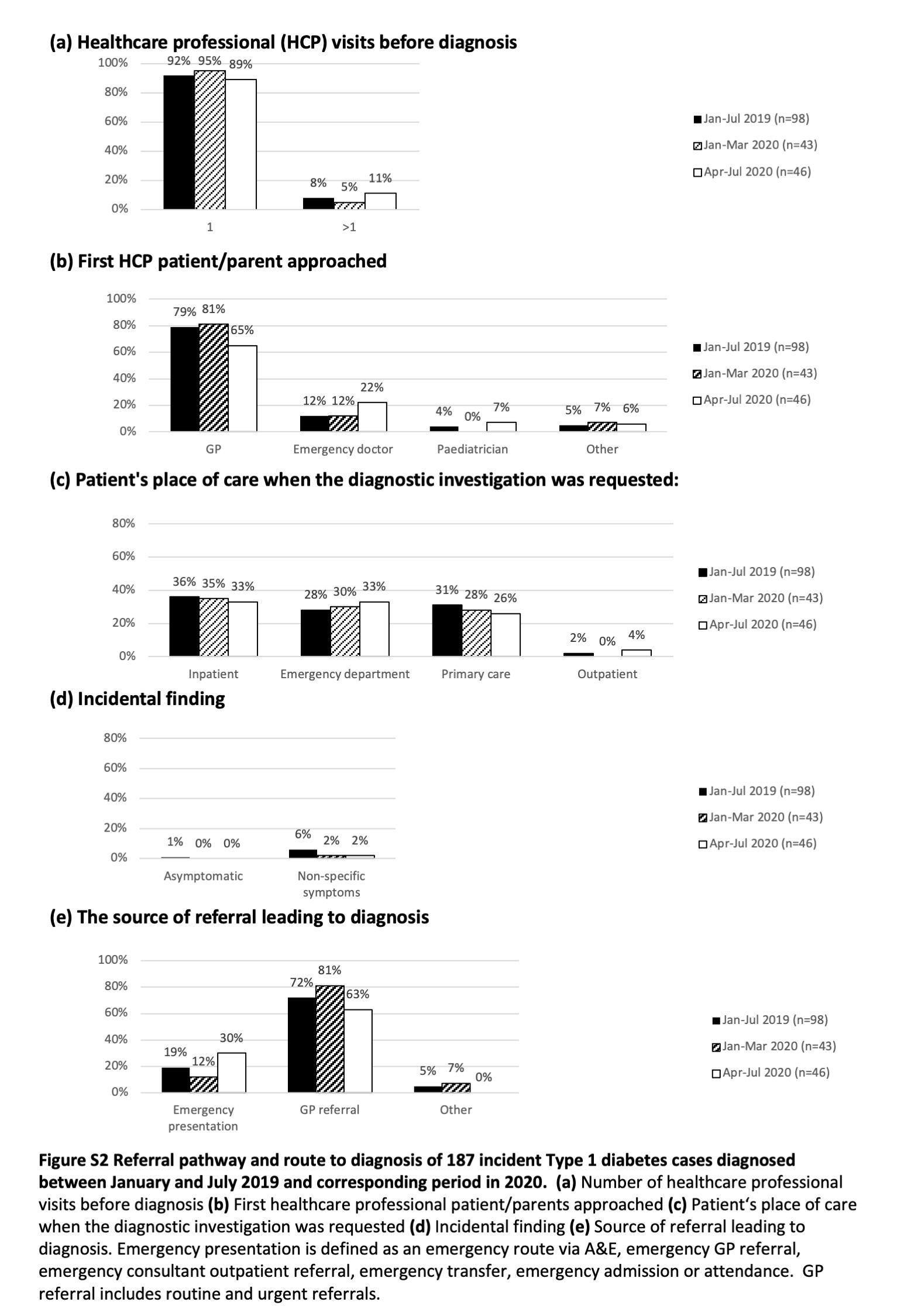

### Figure S3

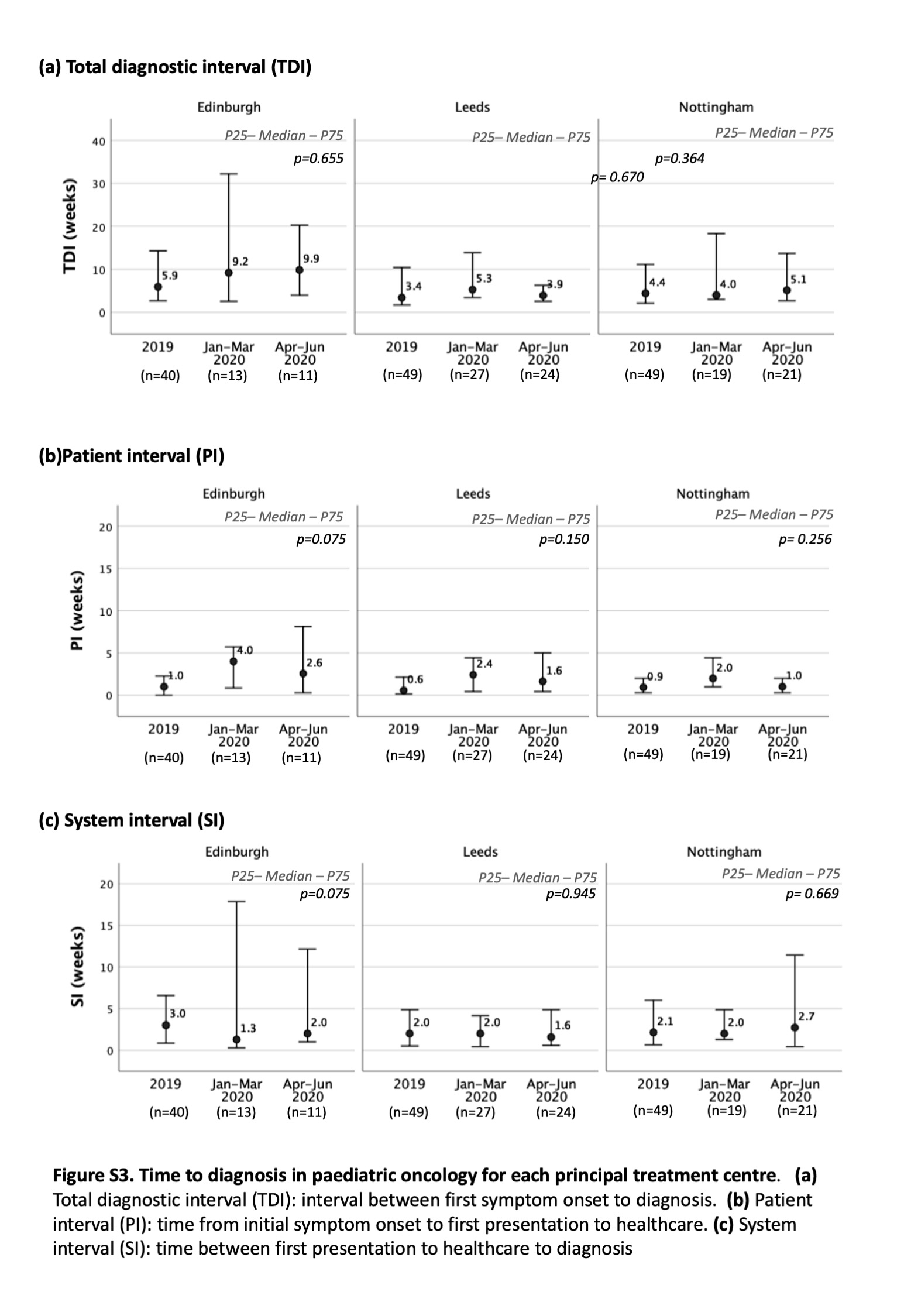

### Figure S4

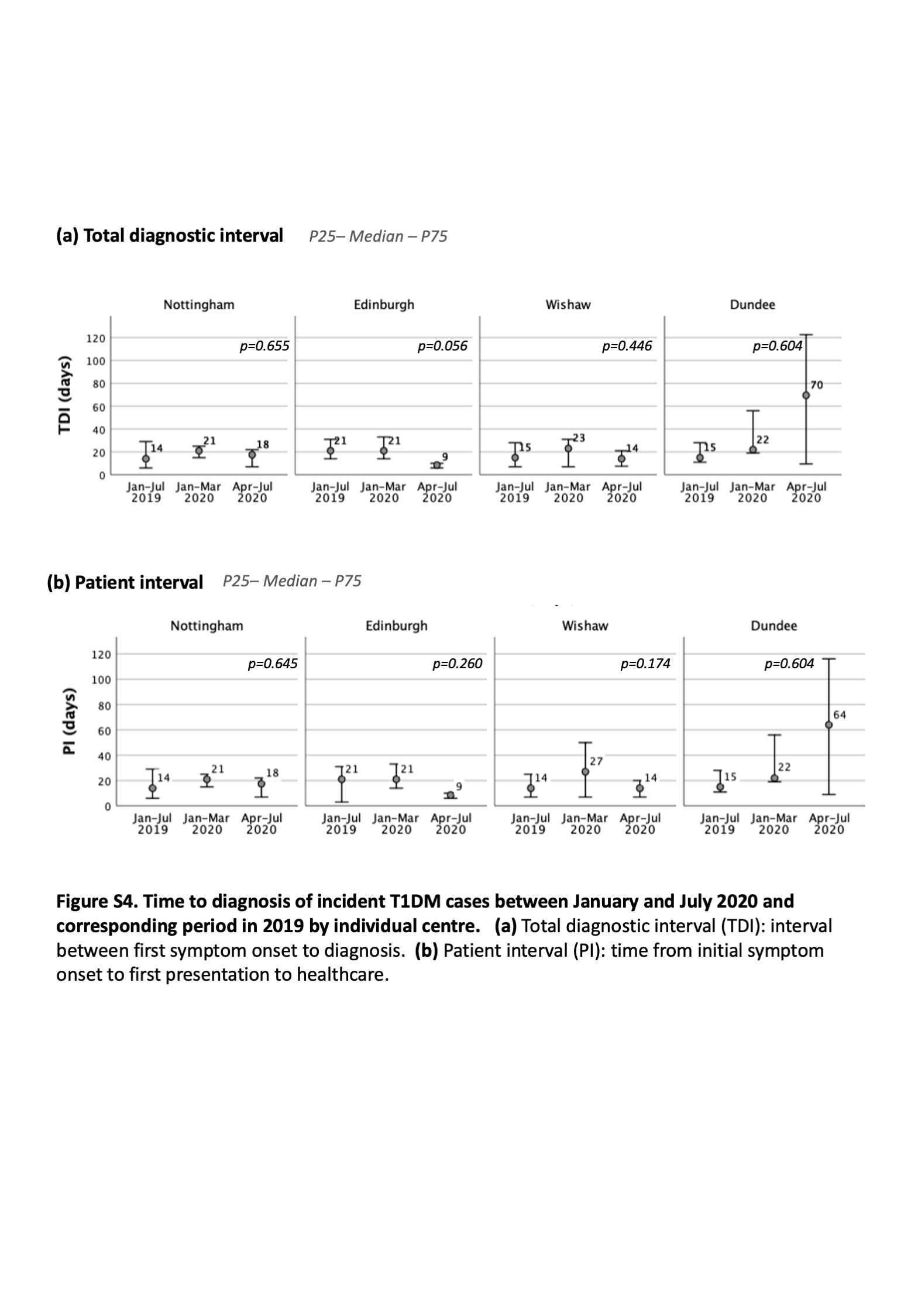
