## Supplementary material for "A multi-centre service evaluation of the impact of the COVID-19 pandemic on presentation of newly diagnosed cancers and type 1 diabetes in children in the UK": Online supplementary material - CC data collection form

### **COVID Pandemic study of urgent clinical presentations of childhood malignancy and new cases of childhood diabetes to Children’s Hospitals / Emergency Departments in UK: A Multicentre Quality Improvement project to inform public and professional guidance for children presenting with serious health concerns at the primary secondary care interface**

**Paediatric Oncology**

**Centre**: _______________________	**Gender**: ☐ Male ☐ Female	**Date of admission:** ______________	**Age at diagnosis:** ☐ Under 5	☐ 5-11 ☐ 12 +

**Clinical Diagnosis:** _____________________	**Patient from a BAME background:** ☐ Yes ☐No

**At symptom onset**

- **Initial symptom(s):** ______________________________________________________________________________

______________________________________________________________________________

- **Key dates (DD/MM/YYYY)**

Date of first symptom onset: ______________________________	☐ Not known

Date of first presentation to healthcare: _____________________	☐ Not known

***Establishing the exact date could be difficult. In this situation please approximate as described below:***

- *If the exact date can only be specified to the nearest week, please record this by entering the date of the first day of the week (Monday).*
- *If the exact date can only be specified to the nearest month, please record this by entering the first day of the month.*
- *If the exact date can only be specified to the nearest season, please record this by entering the first day of April for “spring”, July for “summer” or “mid-year”, October for “fall” or “autumn”. In winter, attempt to determine whether the diagnosis was “late in the year” (use December with the applicable year) or “early in year” (use January with the respective year).*
- *If the information is not available, please select ‘not known*

**Route to diagnosis**

- Was the patient in COVID-19 self-isolation/shielding?

☐ No

☐ Yes	☐ Confirmed case

☐ Possible contact with a confirmed case

☐ Clinical symptoms met

☐ Other ____________________

- Who was the first healthcare professional (HCP) they contacted about these symptoms:

☐ GP	☐ Paediatric emergency doctor	☐ Paediatrician	☐ Dentist	☐ Pharmacist

☐ Optometrist	☐Nurse practitioner	☐ Health visitor	☐ School nurse

☐ NHS 111	☐ Other (please specify ___________________________)

- How many HCP contacts before diagnosis? __________	or	☐ 0-3	☐ 4-6 ☐ 7-9 ☐10+
- Patient's place of care when the investigation that identified the tumour was requested:

☐ Primary care	☐ Outpatient	☐ Inpatient	☐ A&E	☐ Other _______________

- Was this an incidental finding?

☐ No	☐ Yes - asymptomatic	☐ Yes - with non-specific symptoms

- What was the source of referral leading to diagnosis?

| Emergency presentation (A&E) | ☐ Self-referral ☐ GP referral ☐ Optician referral ☐ Dentist referral ☐ MIU/Walk In Centre ☐ Emergency transfer from another hospital  ☐ NHS 111 ☐ Other HCP (please specify) ______________________________ |
| --- | --- |
| GP referral | ☐ Two week wait ☐ Routine referral ☐ Urgent referral to general paediatrician ☐ Other ________________________________________ |
| Other | ☐ Active surveillance (please specify ____________________________) ☐ Diagnosed by another specialty (e.g. ENT)  ☐ Other ________________ |

**At the time of diagnosis**

***Date of diagnosis:*** *the first date of diagnosis whether clinically or histologically established.  
Please record date of clinical diagnosis, date of imaging or date of biopsy, either from pathology report or MDT meeting.*

- Date of diagnosis: ______________________________

☐ Clinical diagnosis	☐Blood test	☐Imaging ☐Biopsy/surgery ☐ Not known

☐	Other ______________

- Did the patients need to be admitted to ICU to start treatment within 7 days of admission?

☐ No	☐ Yes

**MDT**

- Diagnosis (as agreed during MDT meeting): ________________
- Tumour stage: _____________________
- Tumour size (as recorded in radiology report, largest dimension): ___________________
- Clinical risk group (if applicable): _______________________________
- Date starting treatment: ______________________________	☐ Not known

**Emergency Medicine**

**Set A:**

Number of A&E attendance during the time period

**Set B: Data from Emergency Department (if applicable)**

Once received the patient list from Oncology, please extract the following information of individual patient from A&E database:

- Date of ED visit	____________________
- PEWS at triage	____________________
- Triage category	____________________
- Highest level of care required (ward, HDU, PICU) ____________________
- Disposal ____________________
