## Supplementary material for "A multi-centre service evaluation of the impact of the COVID-19 pandemic on presentation of newly diagnosed cancers and type 1 diabetes in children in the UK": Online supplementary material - T1DM data collection form

### **COVID Pandemic study of urgent clinical presentations of childhood malignancy and new cases of childhood diabetes to Children’s Hospitals / Emergency Departments in UK: A Multicentre Quality Improvement project to inform public and professional guidance for children presenting with serious health concerns at the primary secondary care interface**

**Diabetes Mellitus**

**Centre:**_________________________	**Gender**: ☐ Male ☐ Female  **Age at diagnosis:** ☐ Under 5	☐ 5-11 ☐ 12 +	**Patient from a BAME background:** ☐ Yes ☐No

- **Initial symptom(s)**

☐ polydipsia	☐ polyuria	☐ thirst	☐ lethargy	☐ weight loss

☐ other _______________________________________________________________

- **Key dates (DD/MM/YYYY)**

Date of first symptom onset: __________________________________	☐ Not known

Date of first presentation to healthcare: _________________________	☐ Not known

Date of diagnosis: ___________________________________________	☐ Not known

Date starting insulin treatment: ________________________________	☐ Not known

***Establishing the exact date could be difficult. In this situation please approximate as described:***

- *If the exact date can only be specified to the nearest week, please record this by entering the date of the first day of the week (Monday).*
- *If the exact date can only be specified to the nearest month, please record this by entering the first day of the month.*
- *If the information is not available, please select ‘not known*

**Route to diagnosis**

- Was the patient in COVID-19 self-isolation/shielding?

☐ No

☐ Yes	☐ Confirmed case

☐ Possible contact with a confirmed case

☐ Clinical symptoms met

☐ Other ____________________

- Who was the first healthcare professional (HCP) they contacted about the symptom(s):

☐ GP	☐ Emergency doctor	☐ Paediatrician	☐Nurse practitioner	☐ Health visitor
☐ NHS 111	☐ Other (please specify ____________)

- How many HCP contacts before diagnosis? __________
- Patient's place of care when the definitive test (e.g. capillary/blood glucose) that identified the condition was requested:

☐ Primary care	☐ Outpatient	☐ Inpatient	☐ A&E	☐ Other _______________

- Definitive test: ☐ Lab glucose	☐POCT glucose	☐ Other _________________
  Result: ___________________
- Was this an incidental finding?

☐ No	☐ Yes - asymptomatic	☐ Yes - with non-specific symptoms

- What was the source of referral leading to diagnosis?

| Emergency presentation (A&E) | ☐ Self-referral ☐ GP referral ☐ MIU/Walk In Centre  ☐ Emergency transfer from another hospital  ☐ NHS 111 ☐ Other HCP (please specify) ______________________________ |
| --- | --- |
| GP referral | ☐ Routine referral ☐ Urgent referral to general paediatrician  ☐ Urgent referral to specialist diabetic services  ☐ Other ________________________________________ |
| Other | Please specify: ________________ |

- Any other comments about the patient’s journey to diagnosis

_______________________________________________________________________________

_______________________________________________________________________________

- Diabetic ketoacidosis at presentation

☐ No	☐ Yes

Glucose level: ________________

pH at presentation: ________________

Ketones: ________________

Bicarbonate: ________________

GCS at presentation: ________________

Cerebral oedema: ☐ No	☐ Yes

Fluid resuscitation: ☐ No	☐ Yes	amount:______________

Ventilation: ☐ No	☐ Yes	______________ days

ITU stay: ☐ No	☐ Yes	______________ days

- **Antibodies:**

Islet cell cytoplasmic antibodies (ICA)	☐ Negative ☐ Positive	☐ Not tested

Glutamic acid decarboxylase antibodies (GAD)	☐ Negative ☐ Positive	☐ Not tested

Insulin autoantibodies (IAA)	☐ Negative ☐ Positive	☐ Not tested

Zinc Transporter 8 antibodies (ZnT8)	☐ Negative ☐ Positive	☐ Not tested

Insulinoma-associated-2 autoantibodies (IA-2)	☐ Negative ☐ Positive	☐ Not tested

Other: ____________________

**Emergency Department**

**Set A: Overall DKA%**

- Number of A&E attendance
- Number of DKA cases

**Set B: Individual data**

Once received the patient list from Endocrinology/diabetes, please extract the following information of individual patient from A&E database:

- PEWS at triage
- Triage category
- Highest level of care required (ward, HDU, PICU)
- Disposal
