## Supplementary material for "A multi-centre service evaluation of the impact of the COVID-19 pandemic on presentation of newly diagnosed cancers and type 1 diabetes in children in the UK": Online supplementary material - CC cases

### Incident childhood cancer cases during COVID-19 pandemic
#### Jan-July 2020 vs Jan-July 2019

**17 Centres, 2069 patients**
Aberdeen, Belfast, Birmingham, Bristol, Cambridge, Cardiff, East Midlands, Edinburgh, Glasgow, GOSH, Leeds, Liverpool, Newcastle, RMH, Sheffield, Southamptonm, UCLH.

Newly diagnosed childhood cancer cases

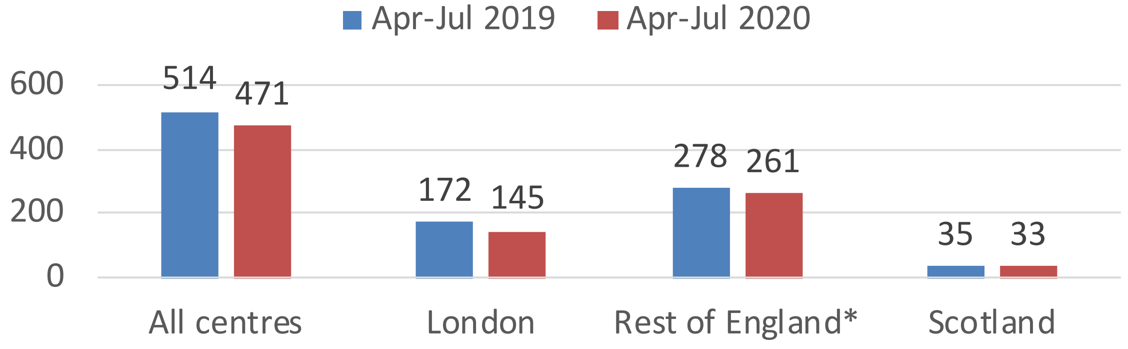


Percentage change
Apr-Jul 2020 vs Apr-Jul 2019

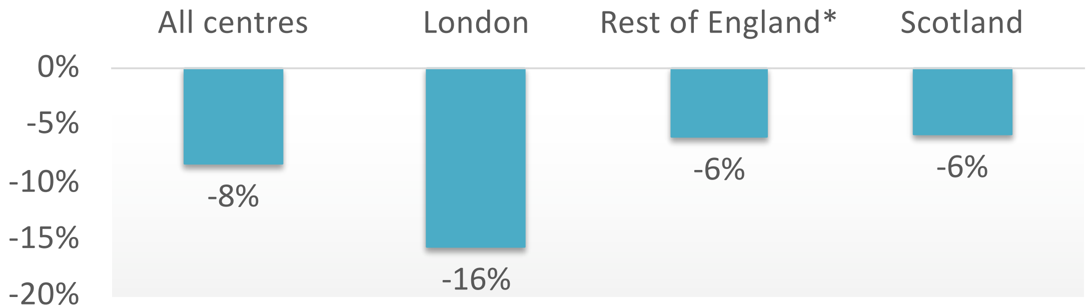


**All 17 centres**

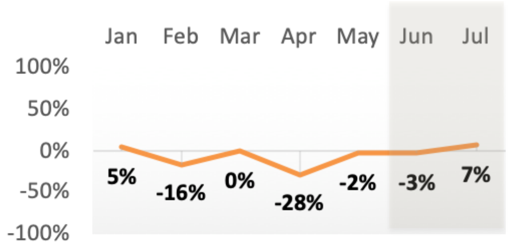


**13 centres with complete data**

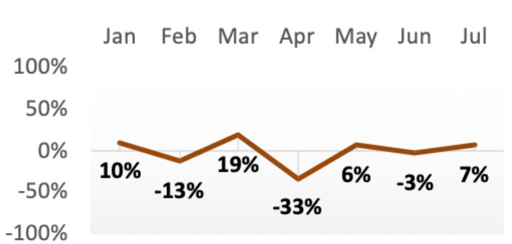


**London**

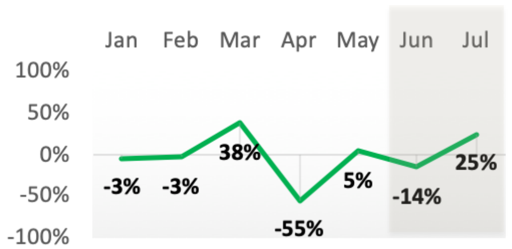


**Rest of England**

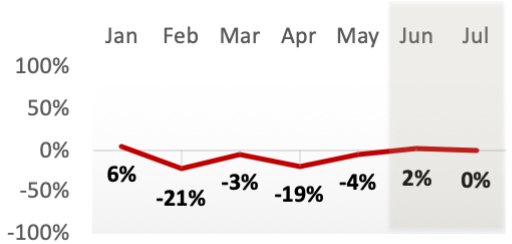


**Scotland**

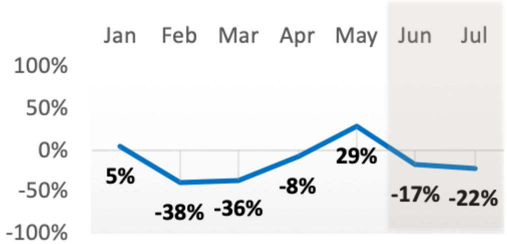


*Shaded areas indicate incomplete data: No June and July data from Birmingham, Cardiff and Southampton*
